# A Retrospective Efficacy and Safety Study of Pembrolizumab/ Cetuximab Neoadjuvant Therapy in Locally Advanced Hypopharyngeal Cancer

**DOI:** 10.1101/2025.02.07.25321054

**Authors:** Guangnan Yao, Xiaobo Wu, Hanqing Lin, Zhihong Chen, Chang Lin, Gongbiao Lin

**Affiliations:** Department of Otolaryngology, Fujian Institute of Otorhinolaryngology, The First Affiliated Hospital of Fujian Medical University, Fuzhou, China; Department of Otolaryngology, Fujian Institute of Otorhinolaryngology, The First Affiliated Hospital of Fujian Medical University, Fuzhou, China Guangnan Yao 、Xiaobo Wu and Hanqing Lin contributed equally to this paper

**Keywords:** hypopharyngeal cancer, neoadjuvant chemotherapy, immunotherapy, tumor response, laryngeal preservation, transoral surgery (TOS)

## Abstract

**Background:** The primary objective of this study was to retrospectively assess the efficacy and safety profiles of two neoadjuvant regimens combining either pembrolizumab or cetuximab with paclitaxel and cisplatin in patients with locally advanced hypopharyngeal cancer (LAHPC).

**Methods:** LAHPC patients who received surgical resection at our hospital between August 2022 and February 2024 were enrolled in the study. All patients received neoadjuvant treatment before surgery and postoperative adjuvant therapy. They were categorized into two groups based on the neoadjuvant regimen: the paclitaxel + cisplatin + pembrolizumab (TP + PEMBRO) group and the paclitaxel + cisplatin + cetuximab (TP + CETUX) group. We evaluated various parameters including treatment response rate, adverse effects, surgical modalities, and survival outcomes for both groups.

**Results:** A total of 32 LAHPC patients were enrolled into the study, with 16 patients in each group. The TP + PEMBRO group demonstrated a significantly superior objective response rate (ORR) of neoadjuvant treatment compared to the TP + CETUX group (87.5% vs 68.75%, P < 0.05). In terms of surgical procedures, the TP + PEMBRO group exhibited a higher proportion of minimally invasive surgeries (87.5% vs 56.25%, P < 0.05), and both the tracheotomy rate and indwelling gastric tube rate were relatively lower in this group. Regarding patient prognosis, the 1-year overall survival (OS) rate in the TP + PEMBRO group was 100%, and the 1-year relapse-free survival (RFS) rate was 92.31%. In contrast, the TP + CETUX group had a 1-year OS rate of 93.75% and a 1-year RFS rate of 81.25%. There was no significant disparity in adverse events between the two groups, and no grade 3 - 4 severe adverse events occurred.

**Conclusion:** The neoadjuvant TP regimen integrating pembrolizumab or cetuximab was associated with higher transoral surgery (TOS) rates and laryngeal preservation rates. Notably, the TP + PEMBRO regimen outperformed the TP + CETUX regimen in terms of treatment response rate and the proportion of minimally invasive surgeries, suggesting a novel and efficacious neoadjuvant treatment for LAHPC.

## 1. Introduction

Hypopharyngeal cancer (HPC) represents a relatively uncommon head and neck malignancy, with 6,475 and 2,314 new cases emerging annually in China and the United States, respectively [1]. The insidious nature of HPC results in over 80% of patients presenting at the locally advanced stage (LAHPC) upon initial diagnosis [2–4]. The prognosis of LAHPC is poor, as the 5-year overall survival (OS) rate hovers around 22 - 30%, and there has been minimal improvement in patient prognosis over the past few decades. Additionally, the recurrence rate of LAHPC is relatively high, with nearly half of patients experiencing recurrence following multimodal treatment [5–7]. Given its proximity to the larynx, the majority of LAHPC patients require total laryngectomy, leading to permanent loss of laryngeal function and severely compromising patients’ quality of life. Therefore, identifying strategies to enhance the prognosis and quality of life of LAHPC patients is an urgent clinical need.

Preoperative neoadjuvant treatment has emerged as a crucial approach, as it can effectively reduce the tumor burden, facilitating preoperative tumor downstaging. This, in turn, can augment the local control rate and overall survival rate, also increasing the likelihood of organ function preservation during surgery. The landmark Veterans Affairs trial and EORTC 24891 trial have firmly established that in locally advanced laryngeal and hypopharyngeal cancers, induction chemotherapy and radiotherapy (RT) can enhance the laryngeal preservation rate without significantly compromising patient prognosis [8,9]. In 2008, the cetuximab-based platinum and fluorouracil regimen (EXTREME regimen) was approved as the first-line treatment for recurrent or metastatic (r/m) head and neck squamous cell carcinoma (HNSCC) [10]. This regimen remained the standard of care for the subsequent decade until the KEYNOTE - 048 and CHECKMATE - 141 studies in 2018 demonstrated the efficacy of PD - 1 immune checkpoint inhibitors in r/m HNSCC [11].

In addition to the EXTREME regimen, paclitaxel combined with cisplatin (TP regimen) is also a commonly employed first-line treatment for LAHPC. Multiple studies have corroborated the safety and efficacy of the TP regimen combined with cetuximab (TP + CETUX) in patients with locally advanced head and neck cancer (LA - HNSCC) [12–14]. Preliminary findings from clinical trials have also suggested that neoadjuvant treatment incorporating PD - 1 inhibitors with TP + CETUX can induce a high pathological tumor regression rate in LA - HNSCC [15–18]. However, to date, no study has directly compared the efficacy of neoadjuvant regimens combining pembrolizumab or cetuximab with TP in LAHPC. This retrospective analysis of LAHPC cases in our center was designed to comprehensively compare the efficacy and safety of these two neoadjuvant regimens.

### 2. Materials and Methods

#### 2.1 Patient Enrollment

This retrospective study encompassed LAHPC patients who visited the Department of Otorhinolaryngology Head and Neck Surgery of the First Affiliated Hospital of Fujian Medical University from August 2022 to February 2024. The inclusion criteria were as follows: 1) Histologically confirmed hypopharyngeal squamous cell carcinoma, with no prior history of anti - tumor treatment; 2) Clinical stage III - IV, and the imaging evaluation indicating a resectable tumor; 3) All patients received neoadjuvant treatment, surgical intervention, and postoperative RT at our center; 4) No distant metastasis detected at the time of initial visit; 5) Patients had regular postoperative follow - up with complete data records available. The exclusion criteria were: 1) A history of other malignancies within the previous 5 years, autoimmune diseases, a history of severe/uncontrolled heart disease, or interstitial lung disease; 2) Previous treatment with immune checkpoint inhibitors; 3) A history of severe infection up to 28 days prior to enrollment.

Patients were stratified into two groups based on the neoadjuvant treatment protocol. One group was administered the pembrolizumab + paclitaxel + cisplatin (TP + PEMBRO) regimen, while the other group received the cetuximab + paclitaxel + cisplatin (TP + CETUX) regimen. This study was approved by the Ethics Committee of the First Affiliated Hospital of Fujian Medical University.

#### 2.2 Treatment Protocols

The treatment strategies for all patients were formulated by a multidisciplinary team (MDT) comprising of medical oncologists, radiation oncologists, pathologists, radiologists, and other specialists.

In the TP + PEMBRO group, patients underwent neoadjuvant treatment with pembrolizumab at a dose of 200 mg on day 1, cisplatin at 75 mg/m² on day 1, and paclitaxel at 175 mg/m² on day 1. Each treatment cycle spanned 21 days, with a maximum of 4 cycles. In the TP + CETUX group, patients received the combination of cisplatin at 75 mg/m² on day 1 and paclitaxel at 175 mg/m² on day 1, along with cetuximab. The initial dose of cetuximab was 400 mg/m², administered via intravenous infusion over 2 hours, followed by a weekly dose of 250 mg/m², infused over 1 hour, for a maximum of 3 cycles.

Regardless of the response of the lesion to neoadjuvant treatment, all patients proceeded to surgical resection. The surgery was scheduled 3 weeks after the completion of the last neoadjuvant treatment cycle. The surgical plan and postoperative follow - up treatment were meticulously determined through MDT discussions. The surgical resection scope was delineated based on post - treatment imaging and electronic laryngoscopy findings. The surgical approach (TOS or open surgery, with or without laryngeal preservation) was selected according to the extent of the lesion. All surgeries were performed by an experienced head and neck surgeon. Lymph node dissection was carried out on the ipsilateral or bilateral neck after neoadjuvant treatment, taking into account the initial metastasis range of LAHPC and the stage of the primary tumor. The margin status of the pathological specimens was analyzed, and histological data were collected to guide adjuvant treatment following MDT consultations.

#### 2.3 Evaluation of Treatment Efficacy and Adverse Reactions

The efficacy of neoadjuvant treatment was evaluated using the Response Evaluation Criteria in Solid Tumors version 1.1. Specifically, by comparing the imaging results after treatment with those at the initial diagnosis, the response to neoadjuvant treatment was categorized as complete response (CR), partial response (PR), stable disease (SD), or progressive disease (PD). Postoperative pathological evaluation was conducted by examining the residual tumor cells in the resected samples. The evaluation of the primary tumor and cervical lymph node (LN) was performed separately. Pathological complete response (PCR) was defined as the absence of any residual tumor tissue in both the primary site and LN metastasis. Major pathological response (MPR) was defined as the presence of less than 10% of viable tumor cells in the primary lesion. Immune partial response (IPR) was defined as a ≥ 30% reduction in the sum of the maximum diameters of all target lesions in the patient, maintained for at least four weeks.

The assessment of adverse events was based on the Common Terminology Criteria for Adverse Events (CTCAE) V5.0. The monitoring and recording of adverse events encompassed the entire neoadjuvant treatment period and extended 30 days after the last neoadjuvant treatment.

#### 2.4 Data Collection and Follow-up

The baseline variables collected in this study included age, gender, smoking history, drinking history, and TNM classification. The treatment - related variables included imaging objective response rate (ORR), surgical method, postoperative pathological complete response rate, tracheotomy, indwelling gastric tube rate and swallowing function, laryngeal preservation rate, short - term survival rate, and adverse events.

Patient follow - up was conducted in accordance with the NCCN guidelines. Specifically, patients were followed up every 2 - 3 months during the first year after surgery and every 4 - 6 months from the second year onwards. Data were collected through outpatient visits or telephone interviews. Additionally, patients underwent electronic laryngoscopy and CT/MR scans during follow - up. The survival outcomes of patients included overall survival (OS) and relapse - free survival (RFS).

#### 2.5 Statistical Analysis

In the statistical analyses, the chi-square test was utilized to compare the baseline characteristics of patients. The Student’s t – test was employed to compare the means between the two groups. The Kaplan - Meier survival curve was used to analyze patient survival. The statistical and graphing software utilized were GraphPad Prism 9 and R 4.1.1. The statistical values were reported as mean ± standard error, and the survival rates were described as 95% confidence intervals. Two-tailed *p* value < 0.05 were defined as statistically significant.

### 3. Results

#### 3.1 Patient Characteristics

A total of 32 male patients were included in this study. The mean age of the patients was 60.41 years (ranging from 49 to 78 years). There were 11 (34.38%) stage III patients and 21 (65.62%) stage IV patients. Nineteen (59.38%) patients had a smoking or drinking history. No significant differences were observed in clinical characteristics such as age, gender, smoking history, drinking history, and tumor stage between the two treatment groups (Table 1).

**Table 1.**
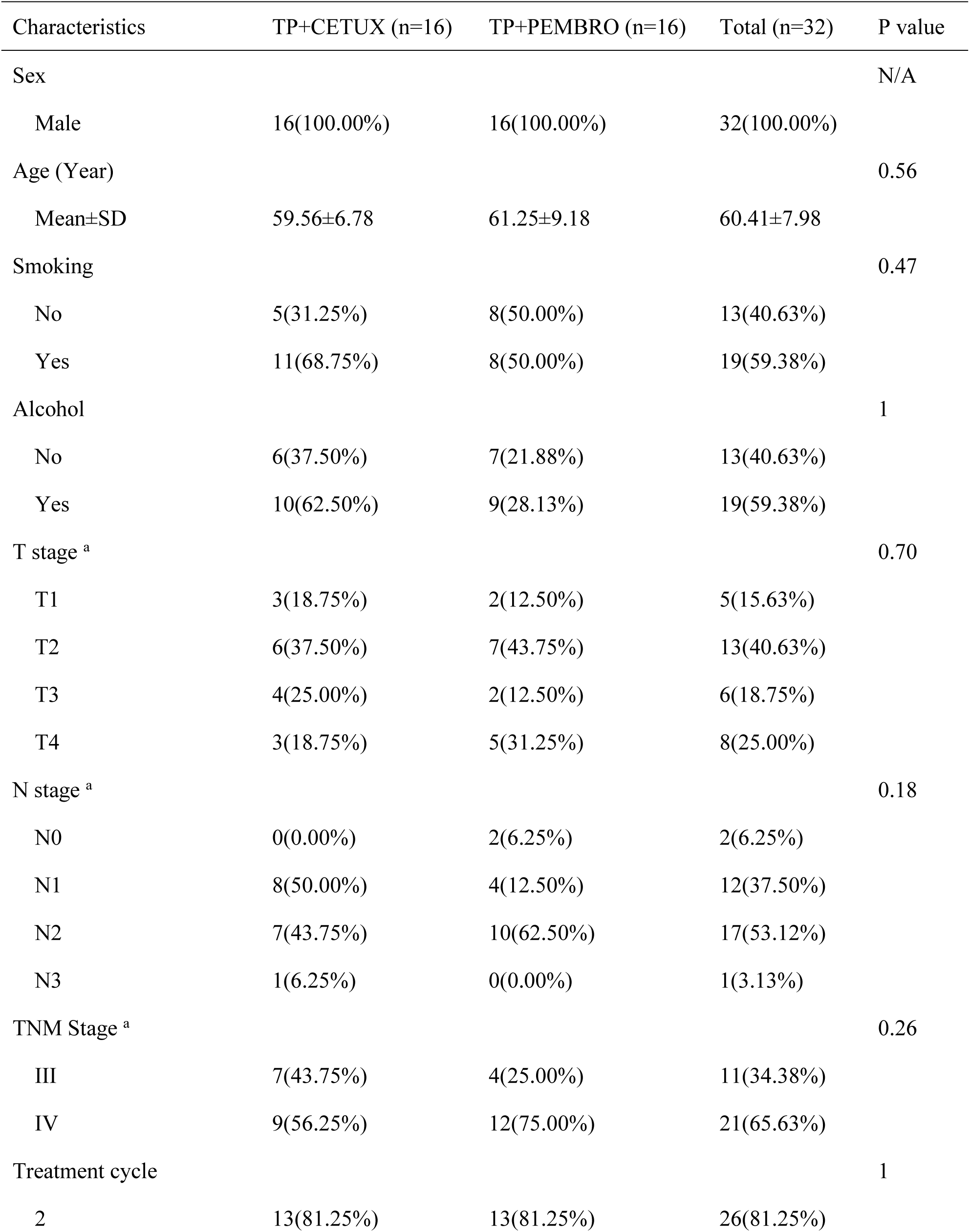
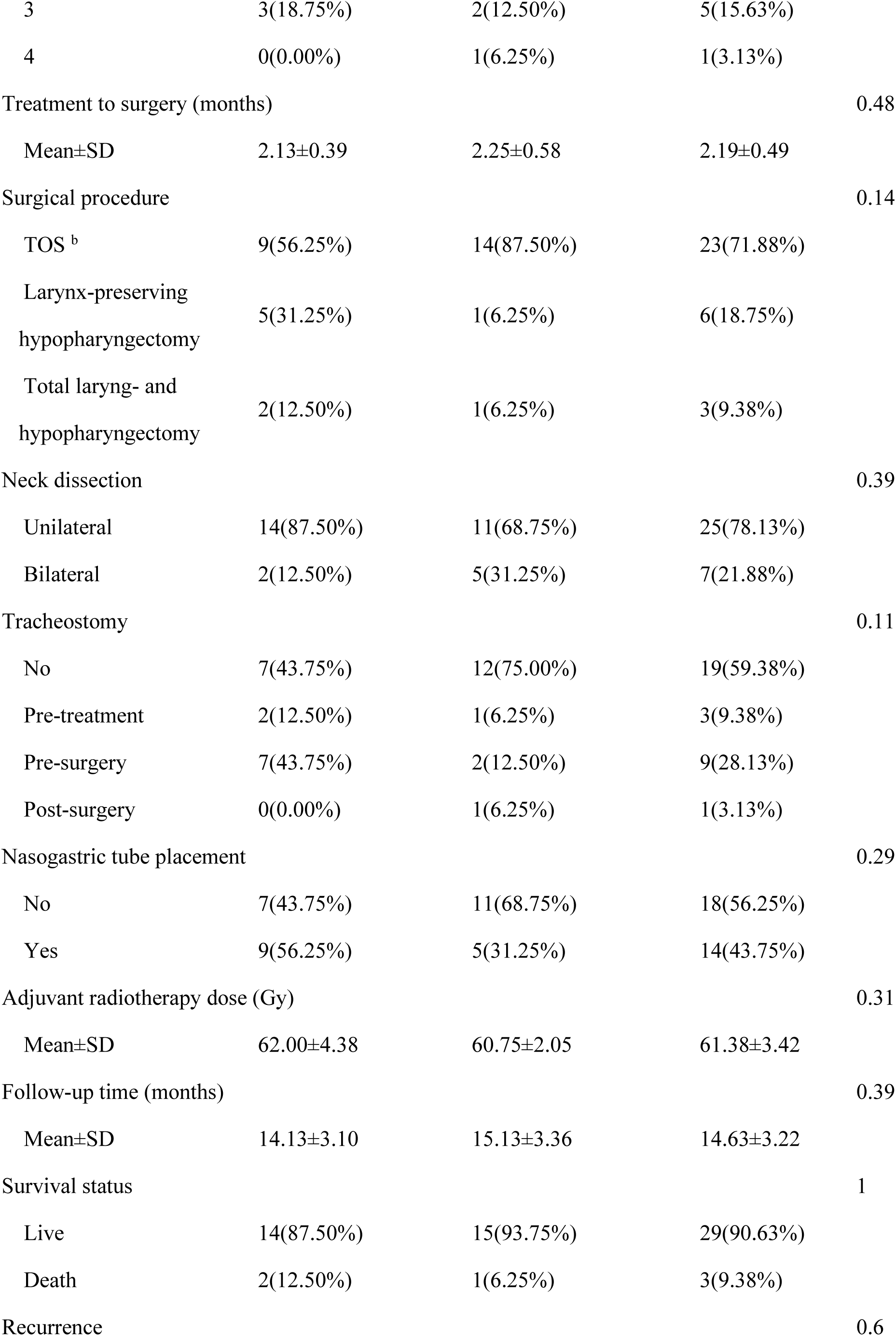
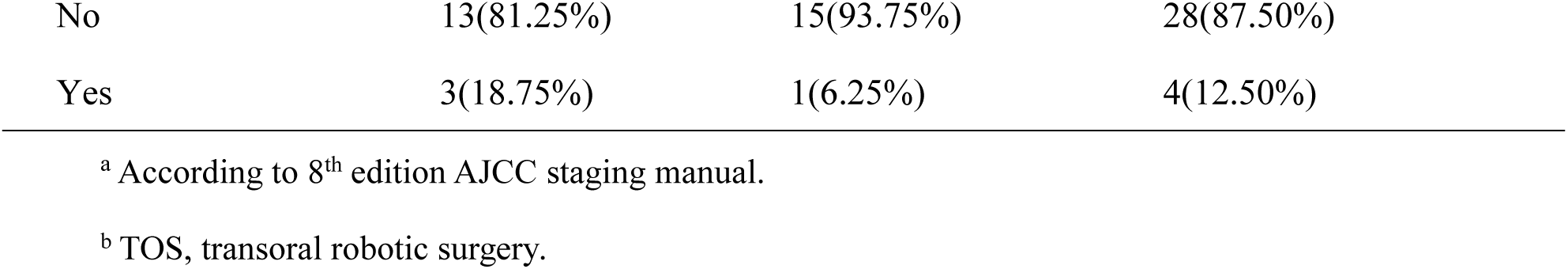
Clinical Characteristics of HPSCC patients

| Characteristics | TP+CETUX (n=16) | TP+PEMBRO (n=16) | Total (n=32) | P value |
| --- | --- | --- | --- | --- |
| Sex |  |  |  | N/A |
| Male | 16(100.00%) | 16(100.00%) | 32(100.00%) |  |
| Age (Year) |  |  |  | 0.56 |
| Mean±SD | 59.56±6.78 | 61.25±9.18 | 60.41±7.98 |  |
| Smoking |  |  |  | 0.47 |
| No | 5(31.25%) | 8(50.00%) | 13(40.63%) |  |
| Yes | 11(68.75%) | 8(50.00%) | 19(59.38%) |  |
| Alcohol |  |  |  | 1 |
| No | 6(37.50%) | 7(21.88%) | 13(40.63%) |  |
| Yes | 10(62.50%) | 9(28.13%) | 19(59.38%) |  |
| T stage <sup>a</sup> |  |  |  | 0.70 |
| T1 | 3(18.75%) | 2(12.50%) | 5(15.63%) |  |
| T2 | 6(37.50%) | 7(43.75%) | 13(40.63%) |  |
| T3 | 4(25.00%) | 2(12.50%) | 6(18.75%) |  |
| T4 | 3(18.75%) | 5(31.25%) | 8(25.00%) |  |
| N stage <sup>a</sup> |  |  |  | 0.18 |
| N0 | 0(0.00%) | 2(6.25%) | 2(6.25%) |  |
| N1 | 8(50.00%) | 4(12.50%) | 12(37.50%) |  |
| N2 | 7(43.75%) | 10(62.50%) | 17(53.12%) |  |
| N3 | 1(6.25%) | 0(0.00%) | 1(3.13%) |  |
| TNM Stage <sup>a</sup> |  |  |  | 0.26 |
| III | 7(43.75%) | 4(25.00%) | 11(34.38%) |  |
| IV | 9(56.25%) | 12(75.00%) | 21(65.63%) |  |
| Treatment cycle |  |  |  | 1 |
| 2 | 13(81.25%) | 13(81.25%) | 26(81.25%) |  |
| 3 | 3(18.75%) | 2(12.50%) | 5(15.63%) |  |
| 4 | 0(0.00%) | 1(6.25%) | 1(3.13%) |  |
| Treatment to surgery (months) |  |  |  | 0.48 |
| Mean±SD | 2.13±0.39 | 2.25±0.58 | 2.19±0.49 |  |
| Surgical procedure |  |  |  | 0.14 |
| TOS<sup>b</sup> | 9(56.25%) | 14(87.50%) | 23(71.88%) |  |
| Larynx-preserving<br>hypopharyngectomy | 5(31.25%) | 1(6.25%) | 6(18.75%) |  |
| Total laryng- and<br>hypopharyngectomy | 2(12.50%) | 1(6.25%) | 3(9.38%) |  |
| Neck dissection |  |  |  | 0.39 |
| Unilateral | 14(87.50%) | 11(68.75%) | 25(78.13%) |  |
| Bilateral | 2(12.50%) | 5(31.25%) | 7(21.88%) |  |
| Tracheostomy |  |  |  | 0.11 |
| No | 7(43.75%) | 12(75.00%) | 19(59.38%) |  |
| Pre-treatment | 2(12.50%) | 1(6.25%) | 3(9.38%) |  |
| Pre-surgery | 7(43.75%) | 2(12.50%) | 9(28.13%) |  |
| Post-surgery | 0(0.00%) | 1(6.25%) | 1(3.13%) |  |
| Nasogastric tube placement |  |  |  | 0.29 |
| No | 7(43.75%) | 11(68.75%) | 18(56.25%) |  |
| Yes | 9(56.25%) | 5(31.25%) | 14(43.75%) |  |
| Adjuvant radiotherapy dose (Gy) |  |  |  | 0.31 |
| Mean±SD | 62.00±4.38 | 60.75±2.05 | 61.38±3.42 |  |
| Follow-up time (months) |  |  |  | 0.39 |
| Mean±SD | 14.13±3.10 | 15.13±3.36 | 14.63±3.22 |  |
| Survival status |  |  |  | 1 |
| Live | 14(87.50%) | 15(93.75%) | 29(90.63%) |  |
| Death | 2(12.50%) | 1(6.25%) | 3(9.38%) |  |
| Recurrence |  |  |  | 0.6 |

|  |  |  |  |
| --- | --- | --- | --- |
| No | 13(81.25%) | 15(93.75%) | 28(87.50%) |
| Yes | 3(18.75%) | 1(6.25%) | 4(12.50%) |

<sup>a</sup> According to 8<sup>th</sup> edition AJCC staging manual.
<sup>b</sup> TOS, transoral robotic surgery.

All patients received 2 to 4 cycles of neoadjuvant treatment. Due to suboptimal response to neoadjuvant treatment, 3 patients in the TP + CETUX group and 2 patients in the TP + PEMBRO group received 3 cycles of neoadjuvant treatment, and 1 patient in the TP + PEMBRO group received 4 cycles of neoadjuvant treatment (P > 0.05).

#### 3.2 Efficacy of Neoadjuvant Treatment and Adverse Events

All patients underwent PET-CT or MRI examinations before and after neoadjuvant treatment to precisely measure the size of the primary tumor and cervical LN and to evaluate treatment efficacy. The overall ORR of all patients was 88.13% (25/32), and no patient exhibited disease progression (PD) (Figure 1A). Notably, in the TP + PEMBRO group, the tumor shrinkage rate was significantly higher than that in the TP + CETUX group (−75.06±27.56% vs -40.72±28.35%, P < 0.01). The ORR proportion in the TP + PEMBRO group was also higher than in the TP + CETUX group (87.5% vs 68.75%, P < 0.05, Figure 1B). Table 2 provides a detailed record of the efficacy evaluation of patients.

**Figure 1.**
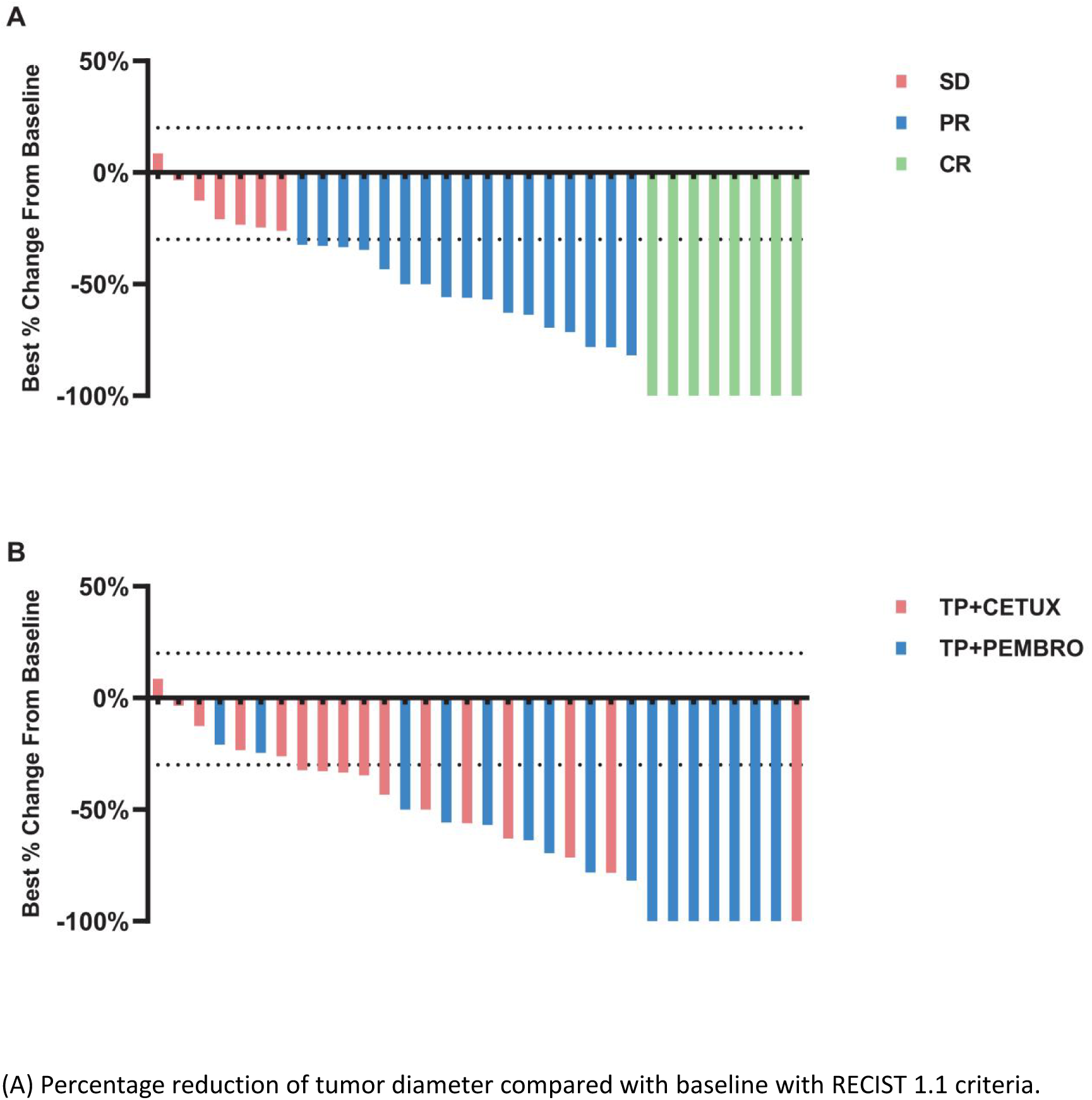
The response of tumors to neoadjuvant therapy. (A) Percentage reduction of tumor diameter compared with baseline with RECIST 1.1 criteria. (B) Tumor regression of patients received different neoadjuvant treatment regimens.

**Table 2.** Response to neoadjuvant chemotherapy

| Characteristics | TP+CETUX (n=16) | TP+PEMBRO (n=16) | Total (n=32) | P value |
| --- | --- | --- | --- | --- |
| Tumor shrinkage (%) |  |  |  | 0.0016* |
| Mean±SD | -40.72±28.35 | -75.06±27.56 | -57.89±32.57 |  |
| Clinical evaluation <sup>a</sup> |  |  |  | 0.04* |
| CR | 1(6.25%) | 7(43.75%) | 8(25.00%) |  |
| PR | 10(62.50%) | 7(43.75%) | 17(53.13%) |  |
| SD | 5(31.25%) | 2(12.50%) | 7(21.88%) |  |
| Total pathologic response <sup>b</sup> |  |  |  | 0.13 |
| IPR | 7(43.75%) | 2(12.50%) | 9(28.13%) |  |
| MPR | 6(37.50%) | 7(43.75%) | 13(40.63%) |  |
| PCR | 3(18.75%) | 7(43.75%) | 10(31.25%) |  |
| Pathologic response (primary tumor) |  |  |  | 0.04* |
| IPR | 7(43.75%) | 2(12.50%) | 9(28.13%) |  |
| MPR | 5(31.25%) | 3(18.75%) | 8(25.00%) |  |
| PCR | 4(25.00%) | 11(68.75%) | 15(46.88%) |  |
| Pathologic response (LN) |  |  |  | 0.0042* |
| IPR | 8(50.00%) | 0(0.00%) | 8(25.00%) |  |
| MPR | 3(18.75%) | 6(37.50%) | 9(28.13%) |  |
| PCR | 5(31.25%) | 10(62.50%) | 15(46.88%) |  |

<sup>a</sup> Evaluated based on RECIST 1.1 criteria. CR, complete response; PR, partial response; SD, stable disease.
<sup>b</sup> Assessed by postoperative pathology. IPR, immune partial response; MPR, major pathologic response; PCR, pathologic complete response.

During neoadjuvant treatment, grade 1 - 2 adverse events were noted among both treatment groups, while no grade 3 - 4 adverse events occurred. The most prevalent adverse events included anemia (62.50%, 20/32), fatigue (56.25%, 18/32), and neutropenia (50.00%, 16/32), among others (Table 3, Figure 2). Throughout the treatment process, no treatment - related adverse events that led to drug discontinuation, dose reduction, or death were observed, nor were any serious immune - related adverse events detected. There was no significant difference in the incidence of adverse events between the two groups treatment groups.

**Table 3.**
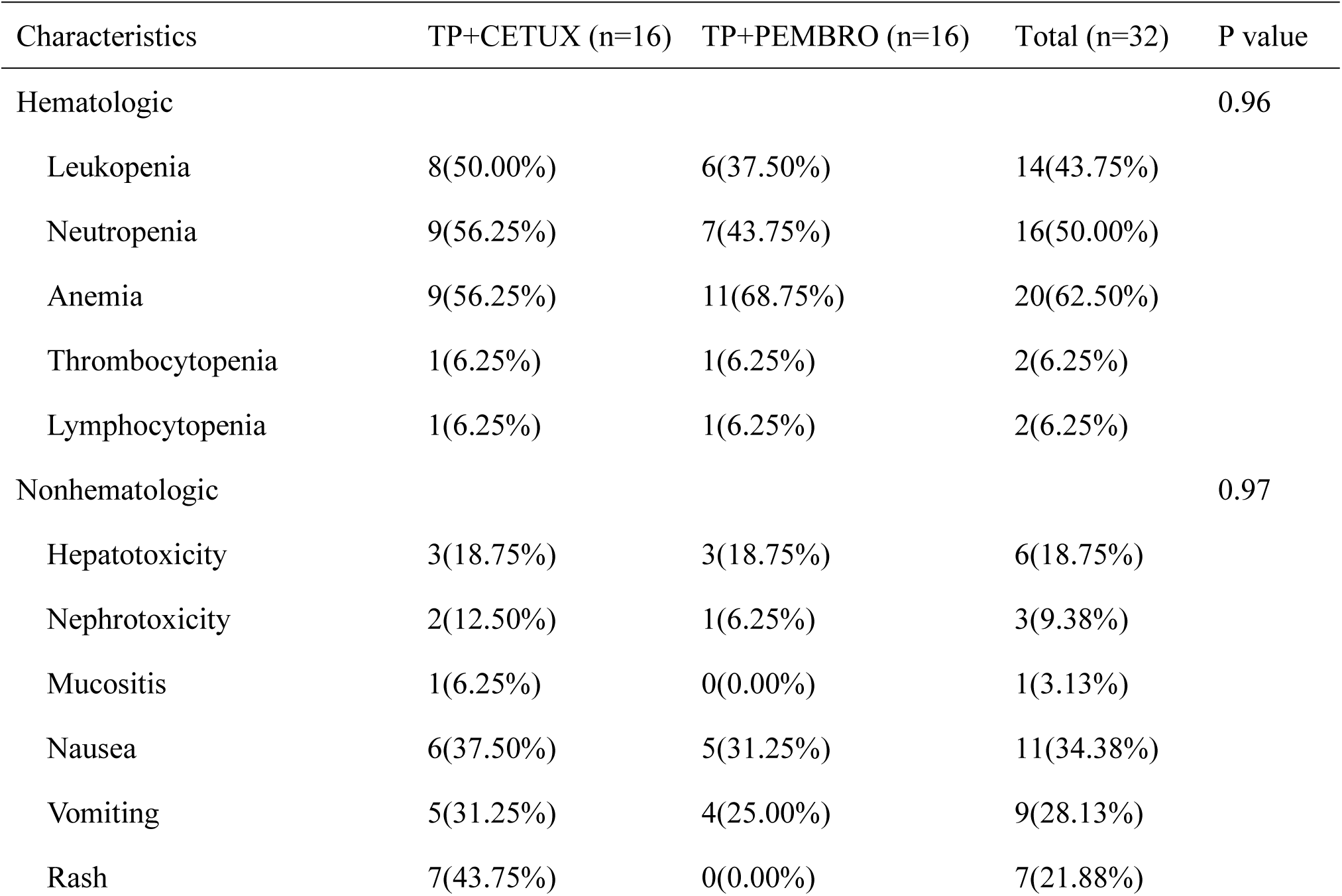
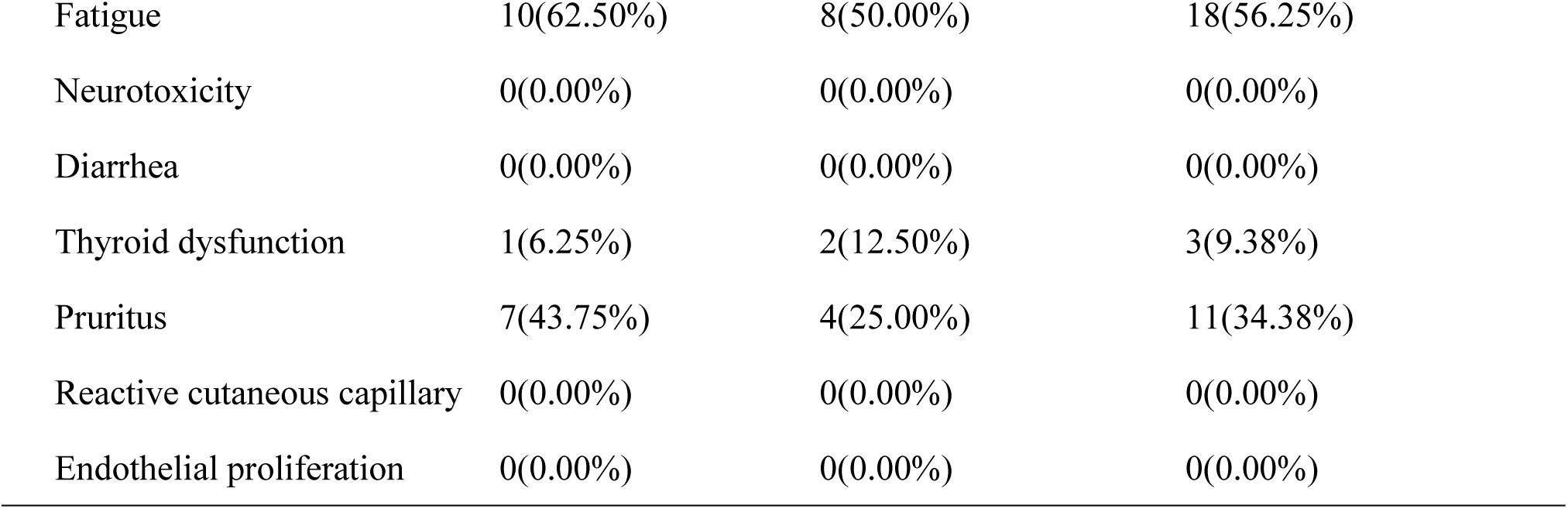
Adverse effects during neoadjuvant treatment

| Characteristics | TP+CETUX (n=16) | TP+PEMBRO (n=16) | Total (n=32) | P value |
| --- | --- | --- | --- | --- |
| Hematologic |  |  |  | 0.96 |
| Leukopenia | 8(50.00%) | 6(37.50%) | 14(43.75%) |  |
| Neutropenia | 9(56.25%) | 7(43.75%) | 16(50.00%) |  |
| Anemia | 9(56.25%) | 11(68.75%) | 20(62.50%) |  |
| Thrombocytopenia | 1(6.25%) | 1(6.25%) | 2(6.25%) |  |
| Lymphocytopenia | 1(6.25%) | 1(6.25%) | 2(6.25%) |  |
| Nonhematologic |  |  |  | 0.97 |
| Hepatotoxicity | 3(18.75%) | 3(18.75%) | 6(18.75%) |  |
| Nephrotoxicity | 2(12.50%) | 1(6.25%) | 3(9.38%) |  |
| Mucositis | 1(6.25%) | 0(0.00%) | 1(3.13%) |  |
| Nausea | 6(37.50%) | 5(31.25%) | 11(34.38%) |  |
| Vomiting | 5(31.25%) | 4(25.00%) | 9(28.13%) |  |
| Rash | 7(43.75%) | 0(0.00%) | 7(21.88%) |  |

|  |  |  |  |
| --- | --- | --- | --- |
| Fatigue | 10(62.50%) | 8(50.00%) | 18(56.25%) |
| Neurotoxicity | 0(0.00%) | 0(0.00%) | 0(0.00%) |
| Diarrhea | 0(0.00%) | 0(0.00%) | 0(0.00%) |
| Thyroid dysfunction | 1(6.25%) | 2(12.50%) | 3(9.38%) |
| Pruritus | 7(43.75%) | 4(25.00%) | 11(34.38%) |
| Reactive cutaneous capillary | 0(0.00%) | 0(0.00%) | 0(0.00%) |
| Endothelial proliferation | 0(0.00%) | 0(0.00%) | 0(0.00%) |

**Figure 2.**
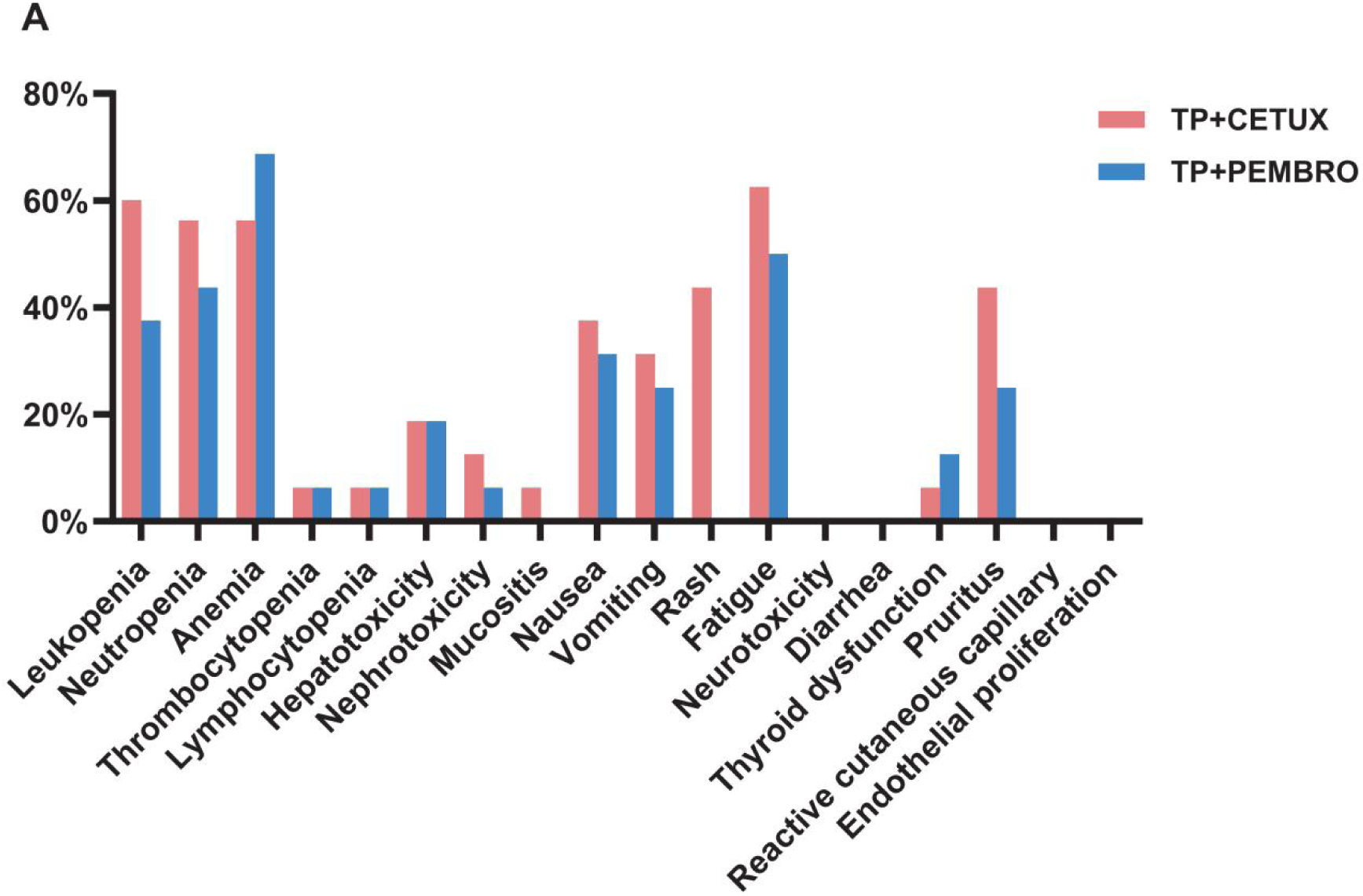
The adverse effects of different neoadjuvant treatment regimens

#### 3.3 Surgical Outcomes

All patients underwent resection of the primary tumor and neck lymph node dissection subsequent to neoadjuvant treatment. Twenty - three patients underwent minimally invasive transoral plasma surgery, while the remaining patients underwent open surgery, with 3 patients undergoing total laryngectomy. The minimally invasive surgery rate in the TP + PEMBRO group was higher than in the TP + CETUX group (87.50% vs 56.25%, P < 0.05), and there was no significant difference in the choice of open surgical methods between the two groups (Table 1).

During the perioperative period, 13 patients underwent tracheotomy, including 4 patients in the TP + PEMBRO group and 9 patients in the TP + CETUX group (Figure 3A). Excluding the patients who underwent total laryngectomy, the average indwelling time of the tracheotomy cannula in the TP + PEMBRO group was 85 ± 30.41 days, while in the TP + CETUX group, it was 171.5 ± 34.10 days (Figure 3B). When compared by surgical method, the average indwelling time of the tracheotomy cannula in the minimally invasive surgery group was 90 ± 34.64 days and 169 ± 34.51 days in the open surgery group(Figure 3C).

**Figure 3.**
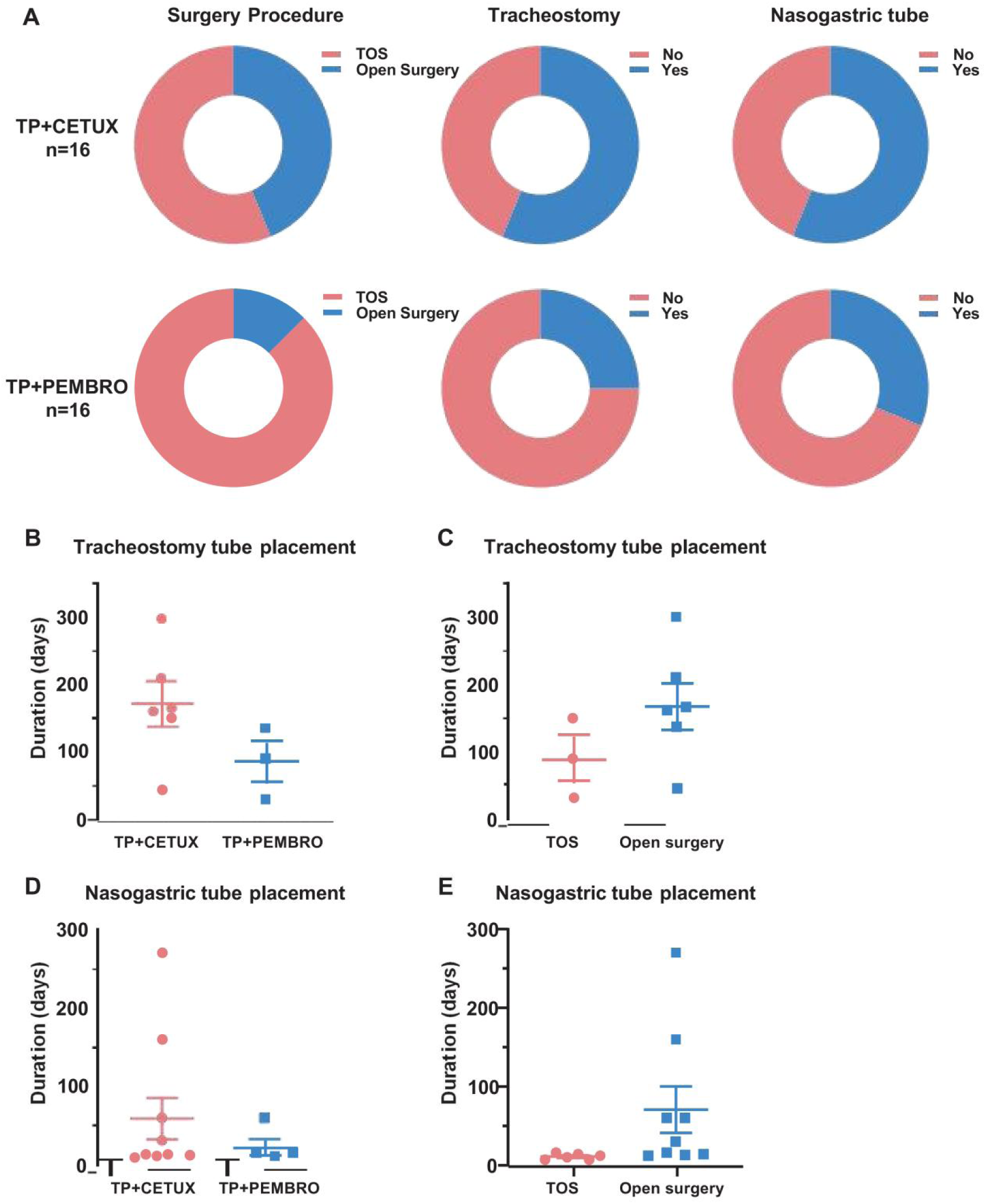
The surgical modalities of patients. (A) The percentage of surgical approach, tracheotomy and indwelling gastric tube in patients received different neoadjuvant regimens. (B-C) The indwelling time of tracheotomy cannula under different neoadjuvant treatment regimens (B) and surgical approaches (C). (D-E) The indwelling time of nasogastric tube under different neoadjuvant treatment regimens (D) and surgical approaches (E).

A total of 14 patients had indwelling gastric tubes for postoperative feeding, with 5 patients in the TP + PEMBRO group and 9 patients in the TP + CETUX group (Figure 3A). The indwelling time of the gastric tube in the TP + PEMBRO group was 22 ± 9.59 days and 59 ± 27.70 days in the TP + CETUX group (Figure 3D). Similarly, when compared by surgical method, the average indwelling time of the gastric tube in the minimally invasive surgery group was 11 ± 1.51 days, and in the open surgery group, it was 70.56 ± 29.48 days (Figure 3E).

#### 3.4 Postoperative Pathological Evaluation

All pathological specimens were evaluated by experienced pathologists after surgery. Overall, 10 patients achieved PCR (31.25%), 13 patients achieved MPR (40.63%), and the remaining 9 patients displayed IPR (28.13%). There was no significant statistical difference in the overall pathological evaluation between the treatment groups (P = 0.13, Table 2).

However, when the primary tumor and LN were evaluated separately, significant differences emerged. In the primary tumor PCR, the rate was 46.88% (15/32), with 68.75% (11/16) in the TP + PEMBRO group and 25.00% (4/16) in the TP + CETUX group; the MPR rate was 25.00% (8/32), with 18.75% (3/16) in the TP + PEMBRO group and 31.25% (5/16) in the TP + CETUX group; the IPR rate was 28.13% (9/32), with 12.50% (2/16) in the TP + PEMBRO group and 43.75% (7/16) in the TP + CETUX group (P value 0.04 < 0.05).

In lymph node PCR, the rate was 46.88% (15/32), with 62.50% (10/16) in the TP + PEMBRO group and 31.25% (5/16) in the TP + CETUX group; the MPR rate was 28.13% (9/32), with 37.50% (6/16) in the TP + PEMBRO group and 18.75% (3/16) in the TP + CETUX group; the IPR rate was 25.00% (9/32), all exhibited in the TP + CETUX group (P value 0.0042 < 0.05) (Table 2).

#### 3.5 Postoperative Adjuvant Treatment and Follow-up

All patients received RT after surgery, with the dose ranging from 50 to 66 Gy. The median follow-up time of patients was 14.63 ± 3.22 months. In terms of patient prognosis, the 1-year OS rate in the TP + PEMBRO group was 100%, and the 1-year RFS rate was 92.31% (95% CI: 56.64 - 98.88%). In the TP + CETUX group, the 1-year OS rate was 93.75% (95% CI: 63.24 - 99.10%), and the 1-year RFS rate was 81.25% (95% CI: 52.46 - 93.54%). There was no significant difference in 1-year OS and RFS between the two groups of patients (Figure 4A - B).

**Figure 4.**
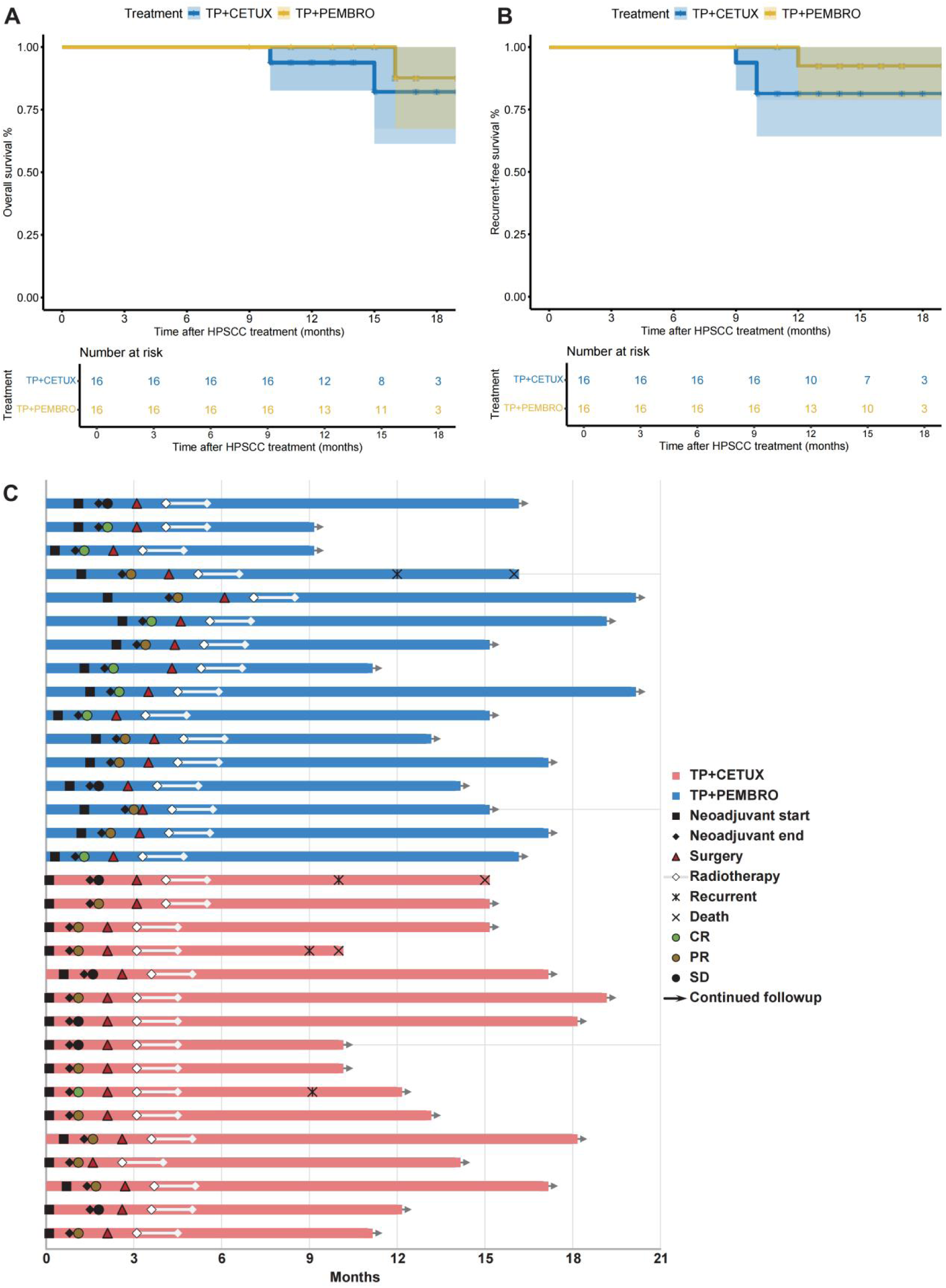
Survival and treatment exposure of patients. (A-B) The overall survival (OS) (A) and relapse-free survival (RFS) (B) of patients. (C) The swimmer plot revealed the treatment exposure and response of neoadjuvant treatment, surgery, and adjuvant therapy in 32 LAHPC patients.

Figure 4C details the main events during the treatment process of all patients. In the TP + PEMBRO group, one patient who passed away had tumor recurrence at the anastomosis 9 months after total laryngectomy and succumbed 13 months after surgery. The postoperative pathological evaluation of this patient was IPR. In the TP + CETUX group, one patient had tumor recurrence 8 months after minimally invasive surgery, with a postoperative pathological evaluation of PCR. This patient underwent total laryngectomy and is still alive as of this writing. Additionally, two patients who underwent open surgery had a postoperative pathological evaluation of IPR and experienced tumor recurrence 7 months after surgery. They died 8 months and 12 months after surgery, respectively.

#### 3.6 A Representative Case

A representative case was one adult patient with hypopharyngeal malignant tumor (T4aN2cM0, IVA)(Figure 5A - B). After two cycles of neoadjuvant treatment with TP + PEMBRO, the tumor regression achieved a complete response (CR)(Figure 5C - D). Subsequently, a transoral surgery (TOS) and bilateral lymph node dissection were performed(Figure 6A). Notably, tracheotomy was not required during the operation. The postoperative pathology confirmed a pathological complete response (PCR). After the operation, the patient received RT. To date, during the 12-month follow-up, the patient has maintained good swallowing and voice functions, and no tumor recurrence has been detected(Figure 6C -D).

**Figure 5.**
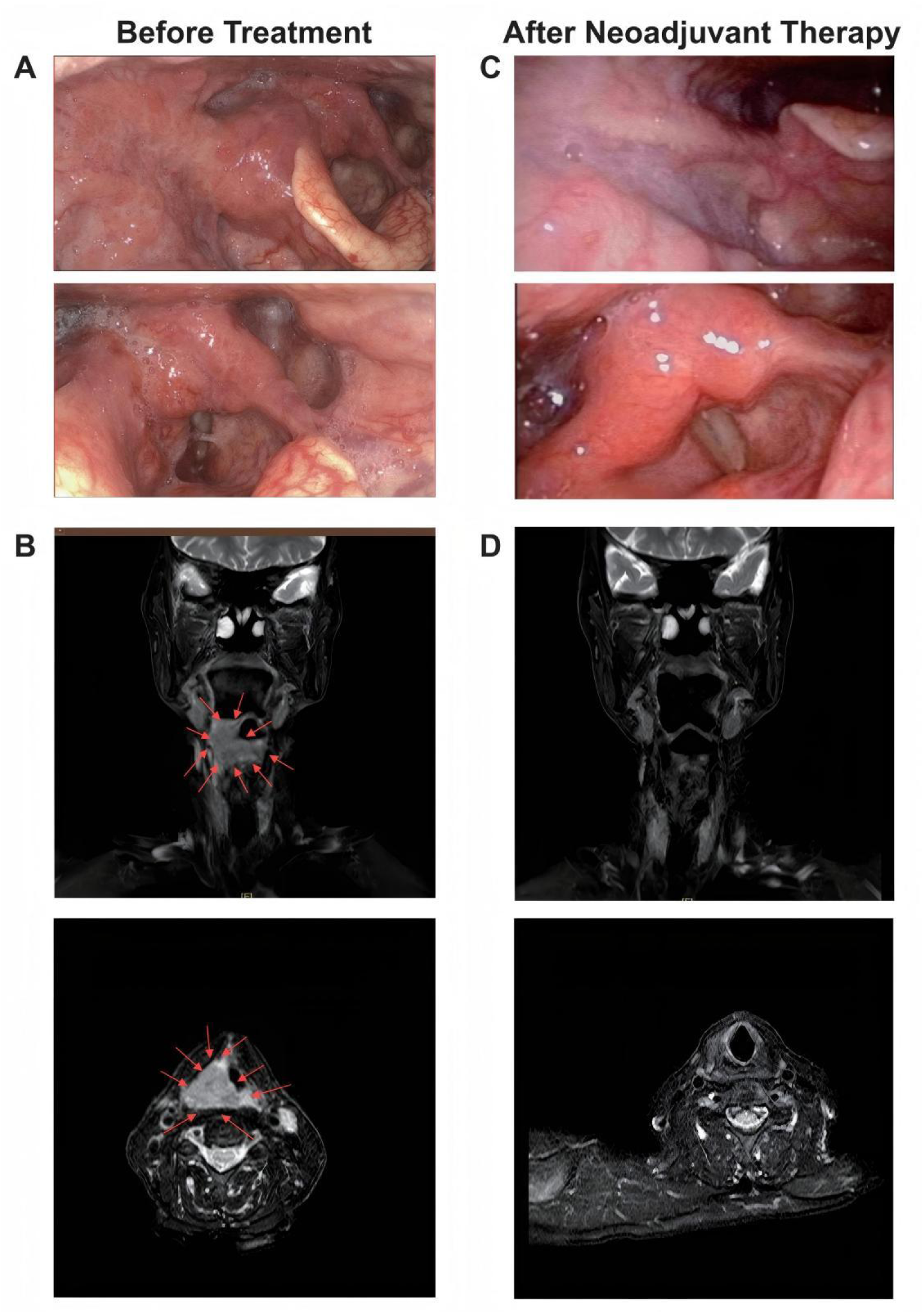
A representative case of tumor regression after neoadjuvant TP+PEMBRO regimen. (A) The scope of the tumor under electronic laryngoscope before neoadjuvant treatment: involving the right lateral wall of the oropharynx, the base of the tongue, the right pyriform sinus, the postcricoid area. (B) The MRI before neoadjuvant treatment. (C) No tumor was found under electronic laryngoscope after neoadjuvant treatment. (D) No tumor was found under MRI after neoadjuvant treatment. The direction of the red arrow indicates the tumor.

**Figure 6.**
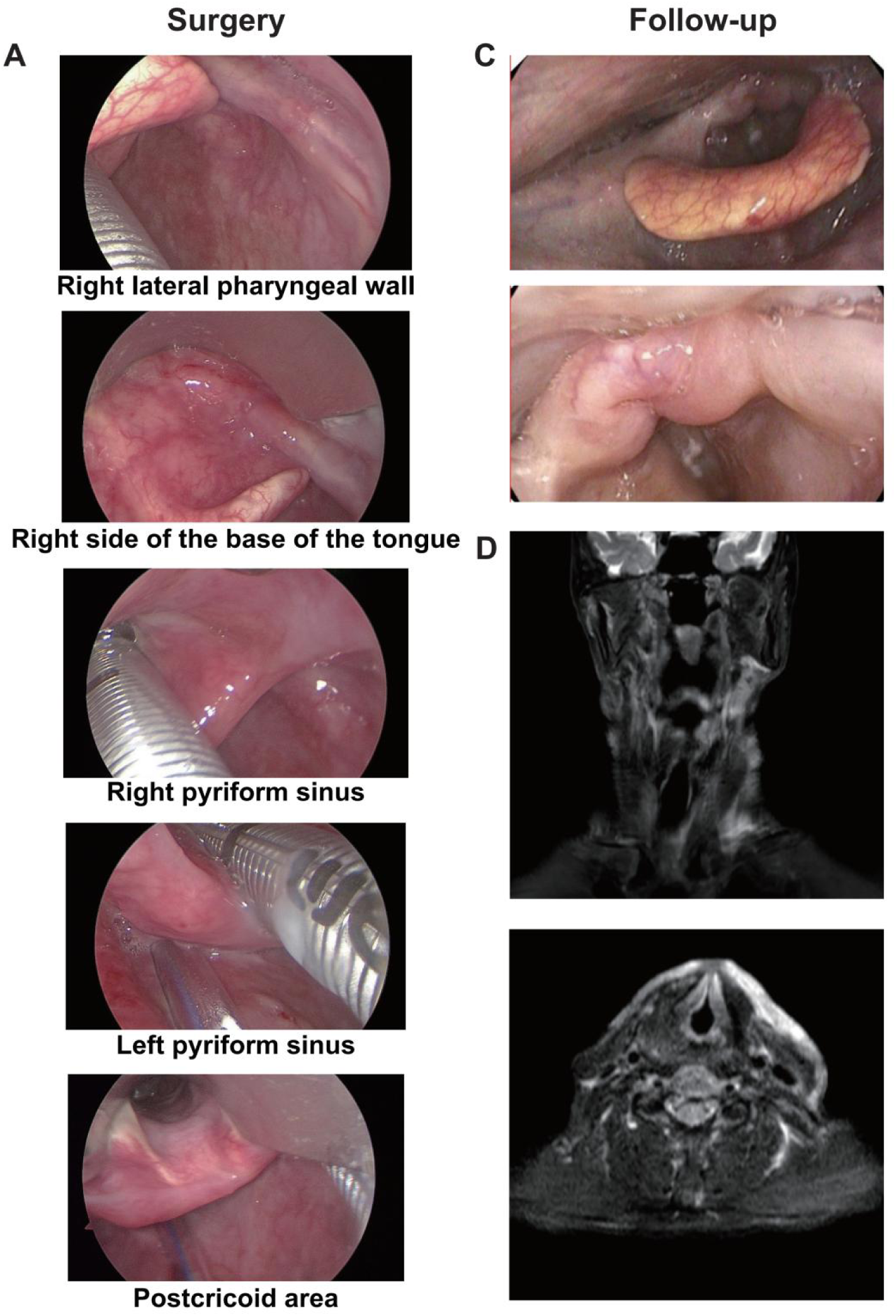
The Intraoperative image and postoperative follow-up of the representative case. (A) The scope of the tumor under suspension laryngoscope endoscopy after neoadjuvant treatment: No tumor was found on the right lateral wall of the oropharynx, the base of the tongue, the right pyriform sinus or the postcricoid area. (C)No tumor was found in the reexamination by electronic laryngoscope one year after the operation. (D)No tumor was found in the reexamination by MRI one year after the operation.

### 4. Discussion

The overarching goal in the management of LAHPC is to optimize function preservation and enhance the quality of life of patients while ensuring survival. Neoadjuvant chemotherapy plays a pivotal role in this context by reducing the tumor burden prior to surgery, thereby potentially increasing the function preservation rate and minimizing the extent of tumor resection [19]. Currently, the conventional neoadjuvant chemotherapy regimens for LAHPC include the TPF regimen and the TP + CETUX regimen. In previous clinical studies, the ORR of these regimens was 63.3% and 74.5%, respectively. However, despite their application, there has been no substantial improvement in patient prognosis [12,14,20,21]. The advent of immunotherapy has introduced novel treatment alternatives for LAHPC patients [22]. In this retrospective study, we analyzed LAHPC patients who received either the neoadjuvant TP + CETUX or TP + PEMBRO regimens and underwent surgical resection at our center. The results demonstrated that the TP + PEMBRO regimen was superior to the TP + CETUX regimen in terms of response rate and minimally invasive surgery rate; it also exhibited a favorable safety profile.

Among the 32 patients included in this study, the overall ORR was 78.13% (25/32). Specifically, the ORR rate in the TP + PEMBRO group was 87.50% (14/16), while in the TP + CETUX group, it was 68.75% (11/16). The response rate of the TP + PEMBRO group was significantly better than that of the TP + CETUX regimen (P < 0.05). No grade 3 or higher adverse events were observed in this study, and the incidence of adverse events in the different treatment groups was comparable, indicating the safety of the neoadjuvant treatment regimens. Previous studies have shown that neoadjuvant chemotherapy combined with PD - 1 inhibitors has demonstrated high pathological remission rates with acceptable safety in LAHNSCC patients [23,24]. The ORR of the TP + CETUX group in this study was consistent with that of previous studies, while the ORR of the TP + PEMBRO group was relatively higher. This discrepancy may be attributed to the significant heterogeneity of HNSCC with different primary sites included in previous studies, as well as differences in drug regimens. These factors make it challenging to accurately evaluate the response and efficacy of neoadjuvant chemotherapy combined with immunotherapy in the treatment of LAHPC. In contrast, all patients in our study were pathologically confirmed LAHPC patients, and the treatment drugs and regimens were relatively uniform, enabling us to precisely assess the response of different treatment regimens.

Reducing the tumor burden, minimizing the tumor resection range, and maximizing the function preservation rate are of utmost importance in neoadjuvant chemotherapy for LAHPC. In this study, 71.88% of patients (23/32) underwent minimally invasive transoral surgery after neoadjuvant treatment. Notably, 87.50% (14/16) of patients in the TP + PEMBRO group underwent minimally invasive surgery, compared to 56.25% (9/16) in the TP + CETUX group (P < 0.05). The overall laryngeal preservation rate of all patients reached 90.62% (29/32), and there was no significant difference in the laryngeal preservation rate between the two groups. Concurrently, in terms of tracheotomy and indwelling gastric tube, 40.62% (13/32) of patients underwent tracheotomy, and 43.75% (14/32) of patients had indwelling gastric tubes during surgery. Although there was no significant difference in the proportion of tracheotomy and indwelling gastric tube and the duration of indwelling between the two groups, the number of cases and the duration of indwelling of tracheotomy and indwelling gastric tube in the TP + CETUX group were generally higher than those in the TP + PEMBRO group. A recent prospective study involving 15 LAHPC patients who received NAC combined with PD - 1 monoclonal antibody treatment reported a total laryngeal preservation rate of 86.6% [25], similar to the laryngeal preservation rate in our cohort. Considering the relatively high proportion of stage IV patients in our cohort (65.62%, 21/32), induction treatment holds significant potential in reducing tumor size, preserving organ functions, and minimizing perioperative complications.

This study also conducted a comprehensive pathological evaluation of patient tumor specimens after surgery. The results revealed that the overall PCR rate was 31.25% (10/32) and the MPR rate was 40.63% (13/32). In the TP + PEMBRO group, the PCR rate was 43.75% (7/16), and the MPR rate was 43.75% (7/16); in the TP + CETUX group, the PCR rate was 18.75% (3/16), and the MPR rate was 37.50% (6/16). Although there was no significant statistical difference in the overall pathological evaluation between the two groups, the number and proportion of PCR and MPR cases in the TP + PEMBRO group were higher than those in the TP + CETUX group. This finding is consistent with previous observations in other tumors, suggesting that immunotherapy combined with neoadjuvant chemotherapy can potentially achieve higher PCR rates than neoadjuvant chemotherapy alone. Additionally, when we evaluated the primary tumor and LN separately, we noted that the pathological remission patterns differed. Fang et al. reported that in locally advanced laryngeal and hypopharyngeal cancers, the response rate of neck lymph node metastases to neoadjuvant chemotherapy combined with immunotherapy was relatively low [26]. However, the results of the CIAO study indicated that the pathological remission effect of lymph node metastases to immune checkpoint inhibitors was better than that of the primary tumor [27]. These findings suggest that there may be heterogeneity in the microenvironment of the primary tumor and lymph node metastases in HNSCC patients. Larger sample clinical studies or multi - omics studies are warranted to further evaluate the correlation and differences in the composition of the microenvironment between different lesions.

For LAHPC patients, current guidelines recommend postoperative adjuvant treatment, which typically includes RT or chemoradiotherapy [28,29]. In this study, all patients received postoperative RT with a dose ranging from 50 to 66 Gy. We also conducted regular follow-up of all patients. The 1-year OS and RFS rates in the TP + PEMBRO group were 100% and 92.31%, respectively; in the TP + CETUX group, the 1-year OS and RFS rates were 93.75% and 81.25%, respectively. During the follow-up process, a total of 4 recurrences were observed, with 3 of these patients eventually dying. The postoperative pathological evaluation of the deceased patients was IPR, while the postoperative evaluation of the surviving patient after recurrence was MPR. This patient remained alive after salvage total laryngectomy. A previous study demonstrated that the 2-year progression - free survival rate of patients who achieved MPR after neoadjuvant immunotherapy was 100%, significantly better than that of IPR patients [30]. However, some studies have also suggested that in HPC, neoadjuvant treatment can improve the laryngeal preservation rate but may not have a significant impact on OS [31,32]. Some researchers have even argued that the current data regarding the impact of tumor response after neoadjuvant immunotherapy on survival remains inconclusive [33]. Although the follow-up period of this study was relatively short, the short - term survival rate of the TP + PEMBRO group was more favorable than that of the TP + CETUX group. Hence, larger sample sizes and longer follow-up durations are essential to further clarify this finding.

Several limitations of the present study must be acknowledged. As a retrospective analysis from a single institution, the small sample size and selection bias cannot be discounted. Additionally, the relatively short follow-up time of the included patients precludes effective evaluation of the long - term efficacy of the treatment. Therefore, future studies should aim to include larger sample sizes and longer follow-up durations to more precisely elucidate the efficacy of different regimens. Furthermore, prospective controlled studies are necessary to r validate the results of this study. Nevertheless, this study has provided preliminary evidence suggesting that the neoadjuvant treatment regimen of TP + PEMBRO may offer greater advantages over the TP + CETUX regimen in LAHPC patients.

### 5. Conclusions

This study demonstrates that both the TP + PEMBRO and TP + CETUX neoadjuvant treatment regimens can effectively preserve the laryngeal function of LAHPC patients and enhance the probability of minimally invasive transoral tumor resection. Moreover, both regimens exhibit high safety profiles. Notably, the TP + PEMBRO group demonstrates certain advantages over the TP + CETUX group in terms of ORR, minimally invasive surgery rate, pathological remission rate, and short - term survival. This study is expected to serve as a valuable theoretical foundation for the future development of neoadjuvant treatment regimens for LAHPC.

## ACKNOWLEDGEMENTS

Not applicable.

## DATA AVAILABILITY STATEMENT

The data sources and handling of the publicly available datasets used in this study are described in the Materials and Methods section.Further details and other data supporting the findings of this study are available from the corresponding author on request.

## FUNDING INFORMATION

Not applicable.

### Disclaimer

All submissions to this journal are the sole responsibility of the authors. The opinions and views expressed in the articles are those of the authors and do not necessarily represent the position of the journal. Authors are responsible for ensuring the originality of their work, obtaining all necessary permissions for any third - party materials used, and disclosing any potential conflicts of interest. The journal does not guarantee the accuracy, completeness, or reliability of the information in the submissions, and shall not be liable for any loss, damage, or liability resulting from the use of the published content.

## ETHICS STATEMENT

The study was approved by the review and ethics committee of The First Affiliated Hospital of Fujian Medical University.

## AUTHOR CONTRIBUTIONS

Guangnan Yao: Data curation; formal analysis; writing original draft.

Xiaobo Wu: Data curation; formal analysis; writing original draft.

Hanqing Lin: Data curation; formal analysis; writing original draft.

Zhihong Chen: curation.

Chang Lin: curation.

Gongbiao Lin: Conceptualization; methodology; supervision.

